# Undiagnosed Dysglycemia and Socioeconomic Status in Argentina: A Paradoxical Gradient in the 2018 National Survey of Risk Factors

**DOI:** 10.64898/2026.03.03.26347343

**Authors:** Matias Agustin Muñoz Nigro

## Abstract

**Background:** Undiagnosed diabetes represents a major challenge for health systems worldwide. While low socioeconomic status is typically associated with reduced healthcare access, the relationship between socioeconomic position and diabetes detection remains poorly characterized in Latin American settings with fragmented health systems.

**Methods:** We analyzed data from 4,409 Argentine adults who underwent capillary glucose measurement in the Third Step of the 2018 National Survey of Risk Factors. Among 471 individuals with elevated glucose (≥110 mg/dL), we examined the association between household income quintile and undiagnosed status using multivariable logistic regression, adjusting for age, sex, health coverage type, education, body mass index, physical activity, and smoking.

**Results:** Contrary to expectations, undiagnosed dysglycemia increased with socioeconomic status: from 45.8% in the lowest quintile to 67.8% in the fourth quintile, with a slight decrease to 61.1% in the highest quintile. After full adjustment, each higher income quintile was associated with 22% greater odds of remaining undiagnosed (OR=1.22; 95% CI: 1.04-1.44; p=0.014). Notably, enrollment in public assistance programs (Plan Estatal) was associated with substantially lower odds of undiagnosed dysglycemia compared to social security coverage (OR=0.27; 95% CI: 0.09-0.79). Results were robust across multiple weighting specifications.

**Conclusions:** Higher socioeconomic status paradoxically increases the likelihood of undiagnosed dysglycemia in Argentina, challenging conventional assumptions about healthcare access. Targeted public programs appear effective at identifying cases among vulnerable populations, while gaps persist in higher-income groups. These findings suggest that diabetes screening strategies should not overlook populations traditionally considered to have adequate healthcare access.

## INTRODUCTION

Diabetes mellitus has emerged as one of the most pressing global health challenges of our time. According to the International Diabetes Federation, approximately 537 million adults worldwide were living with diabetes in 2021, a figure projected to reach 783 million by 2045. Beyond its staggering prevalence, diabetes imposes an enormous burden through microvascular and macrovascular complications, reduced quality of life, and premature mortality.

A particularly troubling aspect of the diabetes epidemic is the substantial proportion of cases that remain undiagnosed. Globally, nearly half of all adults with diabetes are unaware of their condition. This diagnostic gap has profound implications: undiagnosed individuals cannot benefit from interventions that prevent or delay complications, and their disease trajectory often progresses silently until advanced manifestations appear. The consequences extend beyond individual health to encompass increased healthcare costs and lost productivity.

Argentina exemplifies the diabetes challenge facing middle-income countries. National surveys estimate diabetes prevalence at 12.7%, placing it among the highest in Latin America. The Argentine healthcare system presents a distinctive tripartite structure: obras sociales (union-based social security covering formally employed workers), prepaid private insurance, and public sector services that include both the general hospital network and targeted assistance programs for vulnerable populations. This fragmented architecture creates complex patterns of access and utilization that may influence disease detection in ways that differ from more homogeneous systems.

Conventional wisdom, grounded in decades of research from high-income countries, suggests that socioeconomic disadvantage correlates with reduced healthcare access and, consequently, lower rates of disease detection. This pattern aligns with Tudor Hart’s inverse care law, which posits that medical care availability tends to vary inversely with population need, particularly in market-driven systems. However, the applicability of this framework to middle-income settings with mixed public-private healthcare systems remains uncertain.

The 2018 Argentine National Survey of Risk Factors (ENFR) offers a unique opportunity to examine these questions empirically. Unlike previous editions that relied solely on self-reported data, the 2018 survey incorporated a biomarker component (Step 3) with objective glucose measurements in a representative subsample. This design enables identification of individuals with elevated glucose who have never received a diabetes diagnosis the truly undiagnosed population that remains invisible in surveys based only on self-report.

In this study, we investigated the relationship between socioeconomic status and undiagnosed dysglycemia in Argentina, testing whether the expected inverse gradient holds in a fragmented healthcare context. We hypothesized that the distinctive features of the Argentine health system particularly the presence of targeted public programs alongside market-based insurance might produce patterns that diverge from those observed in more homogeneous settings.

## METHODS

### Study Design and Data Source

We conducted a cross-sectional analysis of the 2018 ENFR, a nationally representative household survey administered by Argentina’s National Institute of Statistics and Census (INDEC) in collaboration with the Ministry of Health. The survey employed a probabilistic, multi-stage, stratified cluster sampling design to ensure representation across all Argentine provinces and population strata.

The 2018 ENFR implemented the WHO STEPS methodology, which comprises three sequential levels of data collection. Step 1 gathered information through structured questionnaires on demographics, health behaviors, and self-reported conditions. Step 2 incorporated physical measurements including height, weight, and blood pressure. Step 3, conducted in a random subsample, obtained capillary blood samples for glucose and cholesterol determination. Our analysis focused on participants who completed Step 3.

### Study Population

From 4,409 adults aged 18 years and older with valid glucose measurements, we identified 471 individuals with elevated capillary glucose (≥110 mg/dL). This threshold corresponds to the fasting plasma glucose cut-point of 100 mg/dL recommended for identifying impaired fasting glucose, accounting for the approximately 10% higher values observed in capillary samples. Among these 471 individuals with elevated glucose, we classified 232 (49.3%) as undiagnosed based on negative response to the survey question regarding previous medical diagnosis of diabetes or elevated blood sugar.

### Outcome Definition

The primary outcome was undiagnosed dysglycemia, defined as having elevated capillary glucose (≥110 mg/dL) without prior awareness of this condition. Specifically, individuals were classified as undiagnosed if they had elevated glucose and responded negatively when asked whether a doctor or other health professional had ever told them they had diabetes or elevated blood sugar.

### Exposure Variable

Socioeconomic status was operationalized using household income quintiles derived from self-reported total household income. Quintile 1 represented the lowest-income households and Quintile 5 the highest. This approach captures relative economic position within the Argentine population distribution.

### Covariates

We adjusted for potential confounders based on their established associations with both socioeconomic status and diabetes detection. Demographic variables included age (continuous) and sex (male/female). Health coverage type was categorized as: obra social (union-based social security), prepaid private insurance (prepaga), public assistance program (Plan Estatal), or no formal coverage. Educational attainment served as a secondary socioeconomic indicator. Body mass index was calculated from measured height and weight. Behavioral factors included physical activity level (sufficient vs. insufficient per WHO criteria) and current smoking status.

### Statistical Analysis

We employed survey-weighted logistic regression to account for the complex sampling design. Models were constructed sequentially to examine how the socioeconomic gradient changed with progressive covariate adjustment: Model 1 included only income quintile; Model 2 added age and sex; Model 3 incorporated health coverage type; and Model 4 included the full set of covariates (education, BMI, physical activity, smoking).

To assess potential effect modification by coverage type, we tested the interaction between income quintile and health coverage, followed by stratified analyses within coverage categories. Given concerns about potential variance inflation in complex survey designs, we conducted sensitivity analyses using normalized survey weights, unweighted estimation, and heteroskedasticity-robust standard errors (HC3). All analyses were performed using Python 3.11 with the statsmodels library.

### Ethical Considerations

This study analyzed de-identified public-use data from a government statistical survey. In accordance with Argentine research regulations, secondary analysis of anonymized public-use survey data does not require institutional ethics committee review.

## RESULTS

### Population Characteristics

Table 1 presents characteristics of the study population stratified by glucose status. Among 4,409 participants with glucose measurements, 471 (10.7%) had elevated values. Individuals with elevated glucose were substantially older than those with normal glucose (mean age 55.2 ± 14.9 vs. 45.4 ± 18.0 years, p<0.001), more likely to be male (49.0% vs. 40.6%, p<0.001), and had higher BMI (31.2 ± 6.2 vs. 27.9 ± 5.8 kg/m^2^, p<0.001). Educational attainment was lower in the elevated glucose group, with 15.3% in the lowest education level compared to 7.0% in the normal glucose group, while 43.1% had tertiary education versus 60.2% in the normal group (p<0.001).

**Table 1.** Characteristics of study participants by glucose status, ENFR 2018 Step 3.

| Variable | Normal Glucose<br>(n=3,938) | Elevated Glucose<br>(n=471) | P-value |
| --- | --- | --- | --- |
| Age, years (mean ± SD) | 45.4 ± 18.0 | 55.2 ± 14.9 | <0.001 |
| Female sex, % | 59.4% | 51.0% | <0.001 |
| BMI, kg/m<sup>2</sup> (mean ± SD) | 27.9 ± 5.8 | 31.2 ± 6.2 | <0.001 |
| Current smoker, % | 23.5% | 22.9% | 0.827 |
| Education level, % |  |  | <0.001 |
| Primary or less | 7.0% | 15.3% |  |
| Secondary | 32.7% | 41.6% |  |
| Tertiary/University | 60.2% | 43.1% |  |
| Income quintile, % |  |  | 0.090 |
| Q1 (lowest) | 19.0% | 24.0% |  |
| Q2 | 21.2% | 20.6% |  |
| Q3 | 19.4% | 19.7% |  |
| Q4 | 20.0% | 17.2% |  |
| Q5 (highest) | 20.4% | 18.5% |  |
| Health coverage, % |  |  | 0.031 |
| Obra Social | 63.6% | 69.4% |  |
| Prepaga (private) | 4.6% | 2.3% |  |
| Plan Estatal (public) | 1.4% | 1.9% |  |
| No coverage | 29.6% | 25.7% |  |

*Abbreviations: BMI, body mass index; SD, standard deviation. P-values from t-test (continuous) or chi-square test (categorical).*

### Undiagnosed Dysglycemia by Socioeconomic Status

Among the 471 individuals with elevated glucose, 232 (49.3%) were undiagnosed. The proportion undiagnosed varied markedly across income quintiles, but not in the expected direction. In the lowest income quintile, 45.8% of those with elevated glucose were undiagnosed. This proportion increased progressively to 44.6% in Quintile 2, 51.6% in Quintile 3, and peaked at 67.8% in Quintile 4 before declining slightly to 61.1% in Quintile 5 (Figure 1).

**Figure 1.**
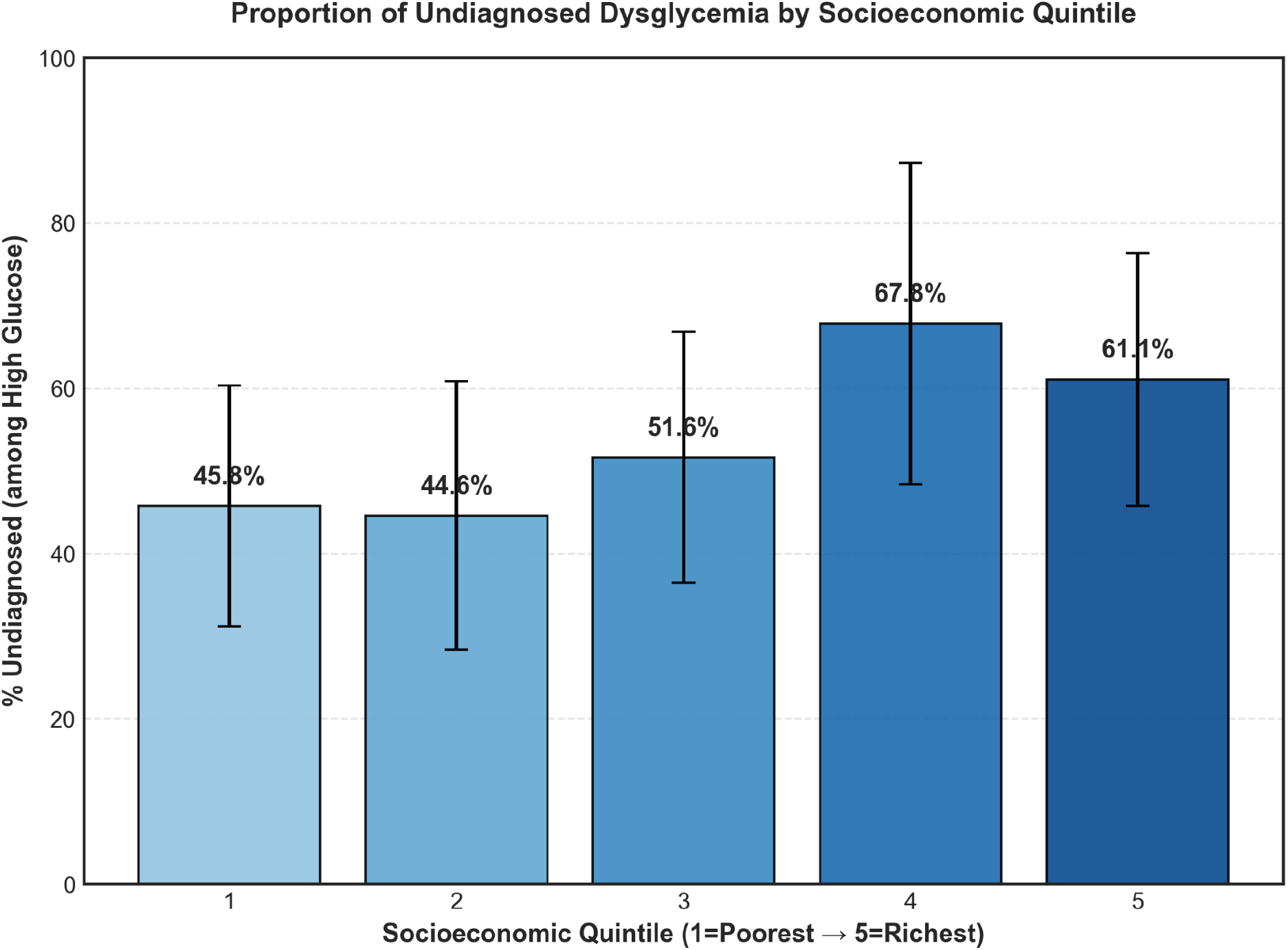
Proportion of individuals with elevated glucose who are undiagnosed, by income quintile. Bars represent weighted proportions; error bars indicate 95% confidence intervals. The paradoxical gradient shows increasing undiagnosed rates with higher socioeconomic status, peaking at Quintile 4 (67.8%).

**Figure 2.**
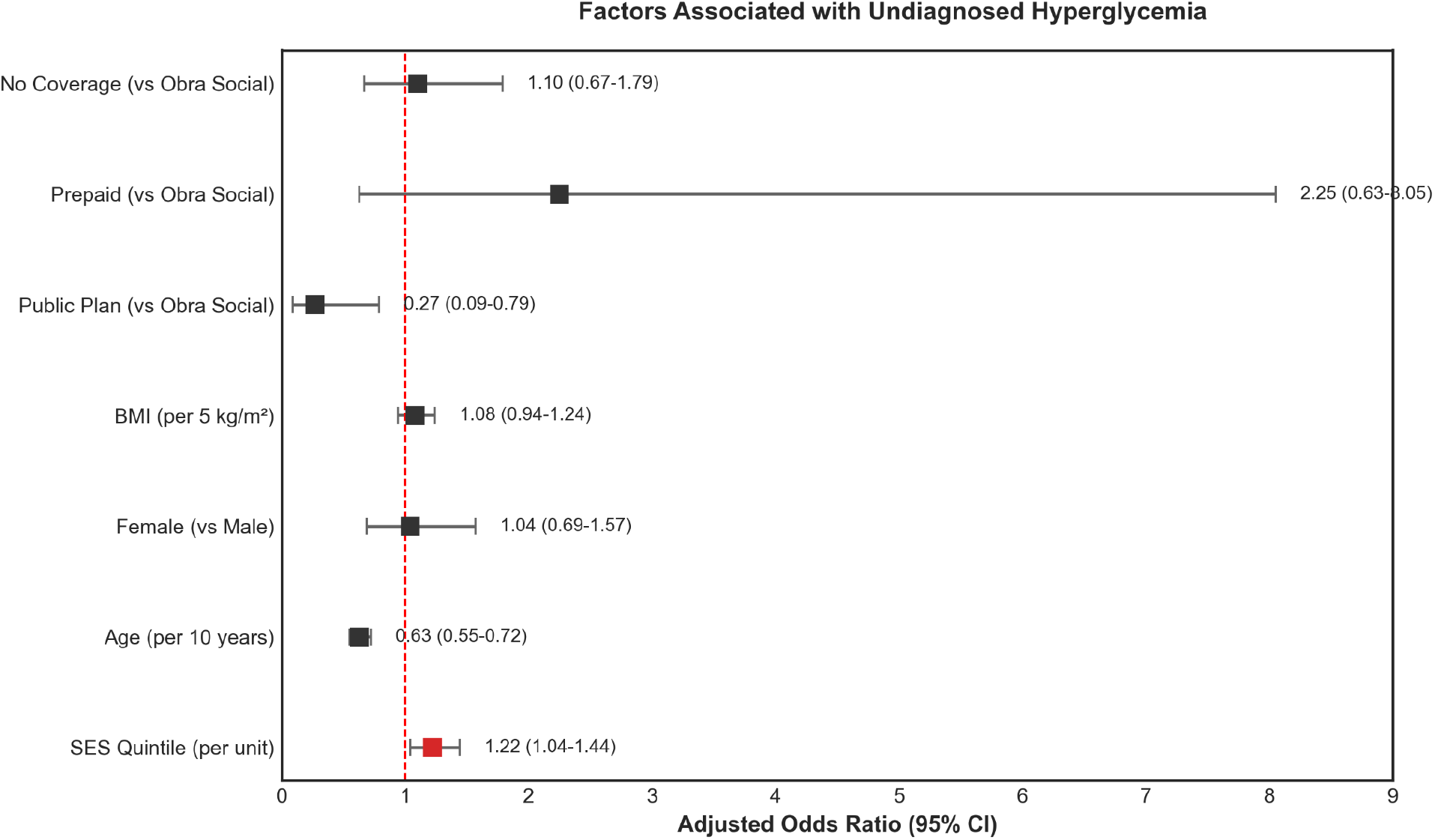
Forest plot of adjusted odds ratios from the fully adjusted model (Model 4). Points represent odds ratios; horizontal lines indicate 95% confidence intervals. The red point highlights the main exposure (income quintile). The vertical dashed line at OR=1.0 indicates no association.

This non-monotonic pattern, with maximum undiagnosed prevalence in the upper-middle income stratum, suggests a detection gap concentrated among individuals with intermediate-to-high socioeconomic status rather than among the poorest populations.

### Multivariable Analysis

Table 2 displays results from sequential logistic regression models. The positive association between income quintile and undiagnosed status was evident in the unadjusted model (Model 1: OR=1.24 per quintile; 95% CI: 1.06-1.44; p=0.006) and remained remarkably stable with progressive covariate adjustment. In the fully adjusted model (Model 4), each higher income quintile was associated with 22% greater odds of being undiagnosed (OR=1.22; 95% CI: 1.04-1.44; p=0.014).

**Table 2.** Multivariable logistic regression for undiagnosed dysglycemia (n=471 with elevated glucose)

| Variable | Model 1 OR (95% CI) | Model 4 OR (95% CI) |
| --- | --- | --- |
| Income quintile (per unit) | 1.24 (1.06-1.44)** | 1.22 (1.04-1.44)* |
| Age (per 10 years) | — | 0.63 (0.55-0.72)*** |
| Female (vs. male) | — | 1.04 (0.69-1.57) |
| BMI (per 5 kg/m<sup>2</sup>) | — | 1.08 (0.94-1.24) |
| Coverage (ref: Obra Social) |  |  |
| Plan Estatal | — | 0.27 (0.09-0.79)* |
| Prepaga | — | 2.25 (0.63-8.05) |
| No coverage | — | 1.10 (0.67-1.79) |

\* $p < 0.05$ ; \*\* $p < 0.01$ ; \*\*\* $p < 0.001$ . Model 1: unadjusted. Model 4: adjusted for age, sex, coverage type, education, BMI, physical activity, and smoking. OR, odds ratio; CI, confidence interval; BMI, body mass index.

Age demonstrated a strong protective association: each 10-year increase in age was associated with 37% lower odds of being undiagnosed (OR=0.63; 95% CI: 0.55-0.72; p<0.001), likely reflecting cumulative opportunities for detection and symptom-driven diagnosis in older individuals. Sex, BMI, physical activity, and smoking did not show statistically significant associations with undiagnosed status after adjustment.

### Health Coverage Type and Detection

The type of health coverage emerged as a powerful predictor of diagnostic status. Compared to obra social (the reference category), individuals enrolled in Plan Estatal (public assistance programs) had substantially lower odds of undiagnosed dysglycemia (OR=0.27; 95% CI: 0.09-0.79; p=0.017). This finding indicates that targeted public programs are effectively reaching and screening vulnerable populations. In contrast, those with prepaid private insurance showed a trend toward higher odds of remaining undiagnosed (OR=2.25; 95% CI: 0.63-8.05), though this estimate was imprecise due to small sample size (n=11 in this category). Individuals without any formal coverage showed odds similar to those with obra social (OR=1.10; 95% CI: 0.67-1.79).

### Interaction and Stratified Analyses

The interaction between income quintile and health coverage type was statistically significant (p<0.001), indicating that the socioeconomic gradient in undiagnosed dysglycemia varied across coverage categories. In stratified analyses, the positive association between income and undiagnosed status was strongest among those without formal coverage (OR=1.59 per quintile) and weakest among those enrolled in public programs (OR=1.17). This pattern suggests that public assistance programs partially attenuate the paradoxical gradient observed in the overall population.

### Sensitivity Analyses

Results were robust across alternative analytic specifications (Supplementary Figure 1). The odds ratio for income quintile ranged from 1.18 to 1.22 across four approaches: original survey weights (OR=1.22), normalized weights (OR=1.22), unweighted analysis (OR=1.18), and heteroskedasticity-robust standard errors (OR=1.18). In all specifications, confidence intervals excluded 1.0 and p-values remained below 0.05, confirming the robustness of our findings.

## DISCUSSION

This nationally representative study reveals a paradoxical pattern in diabetes detection: contrary to conventional expectations, higher socioeconomic status was associated with greater likelihood of undiagnosed dysglycemia in Argentina. This finding challenges assumptions derived from high-income country contexts and highlights the complex dynamics of healthcare access in fragmented health systems.

### A Reversed Gradient

The observed pattern with undiagnosed rates rising from 46% in the lowest income quintile to 68% in the fourth quintile represents an inversion of what Tudor Hart’s inverse care law would predict. In classical formulations, populations with greatest need (typically those of lower socioeconomic status) receive least care. Our findings suggest that in Argentina’s tripartite healthcare system, this relationship may operate differently for preventive services like diabetes screening.

Several mechanisms might explain this counterintuitive gradient. First, targeted public health programs appear remarkably effective at reaching low-income populations. The 73% reduction in undiagnosed odds associated with Plan Estatal coverage (OR=0.27) demonstrates that deliberate policy interventions can overcome structural barriers to detection. These programs often incorporate routine screening protocols as part of comprehensive primary care, ensuring that vulnerable individuals encounter diagnostic opportunities even without presenting symptoms.

Second, the private and obra social sectors may prioritize acute and curative services over preventive care. Higher-income individuals with obra social or prepaid coverage might experience fragmented care with limited emphasis on population-level screening. When these individuals do access care, it may be for specific complaints rather than comprehensive health assessments that would include glucose testing.

Third, symptom-driven diagnosis may be more common in lower socioeconomic groups. When individuals from disadvantaged backgrounds finally present to care often at more advanced disease stages they may be more likely to have overt hyperglycemia that prompts immediate diagnosis. Conversely, higher-income individuals with milder, asymptomatic elevation may never undergo testing despite having nominally better access.

### The Non-Monotonic Pattern

The peak in undiagnosed prevalence at Quintile 4 rather than Quintile 5 suggests additional complexity. The slight decline in the highest income quintile might reflect that the most affluent individuals who can afford comprehensive private care do receive adequate preventive services. The “detection gap” appears concentrated in the upper-middle class: individuals wealthy enough to be excluded from targeted public programs but not wealthy enough (or sufficiently motivated) to seek comprehensive preventive care privately.

### Contextual Considerations

Our findings resonate with emerging evidence from other Latin American contexts. Brazil’s universal health system has achieved considerable success in diabetes care among lower socioeconomic groups, while studies from Colombia and Mexico document persistent detection gaps despite expanding coverage. The common thread appears to be that universal coverage alone is insufficient; targeted, proactive screening programs are necessary to ensure detection across the socioeconomic spectrum.

The Argentine context offers particular insight into how health system architecture shapes these patterns. Unlike systems with unified financing or single-payer models, Argentina’s fragmented structure creates distinct pathways to care with varying emphasis on prevention. The success of Plan Estatal programs demonstrates that within this fragmented landscape, intentional policy design can achieve equitable detection perhaps more effectively than market-based mechanisms.

### Policy Implications

These findings carry several implications for diabetes prevention and control strategies. First, screening programs should not assume that higher-income populations have adequate access to preventive services. Our data suggest the opposite may be true, at least in fragmented health systems. Second, the effectiveness of Plan Estatal programs merits attention and potential expansion. Understanding and replicating the mechanisms through which these programs achieve superior detection rates could inform health policy more broadly. Third, the obra social and prepaid sectors might benefit from strengthened incentives or requirements for preventive screening, perhaps through quality metrics or coverage mandates.

### Strengths and Limitations

This study benefits from several methodological strengths. The ENFR provides nationally representative data with rigorous sampling procedures. The incorporation of objective glucose measurements enables identification of truly undiagnosed cases invisible to self-report surveys. Our analytic approach with sequential models illuminates how the association evolves with covariate adjustment, while sensitivity analyses confirm robustness.

Several limitations warrant acknowledgment. The cross-sectional design precludes causal inference; we cannot determine whether socioeconomic status influences detection or whether other unmeasured factors explain the observed associations. The small number of individuals with prepaid coverage (n=11 with elevated glucose) limits precision for this subgroup. Single glucose measurements may misclassify some individuals, though this would likely bias toward the null. We could not assess disease duration or severity, which might differ across socioeconomic groups in ways that influence detection. Finally, capillary glucose measurement introduces some imprecision compared to venous sampling, though the standardized protocol and population-level focus mitigate this concern.

## CONCLUSIONS

In Argentina, higher socioeconomic status is paradoxically associated with greater likelihood of undiagnosed dysglycemia, challenging conventional assumptions about healthcare access and disease detection. Targeted public programs demonstrate remarkable effectiveness at reaching vulnerable populations, while detection gaps persist among higher-income groups. These findings suggest that diabetes screening strategies must account for the specific dynamics of fragmented health systems, where nominal access does not guarantee utilization of preventive services. Interventions to improve detection should span the socioeconomic spectrum rather than focusing exclusively on disadvantaged populations.

## Data Availability

The 2018 ENFR data are publicly available from INDEC (Instituto Nacional de Estadistica y Censos de Argentina) at https://www.indec.gob.ar/.

## FIGURE LEGENDS

**Supplementary Figure 1.**
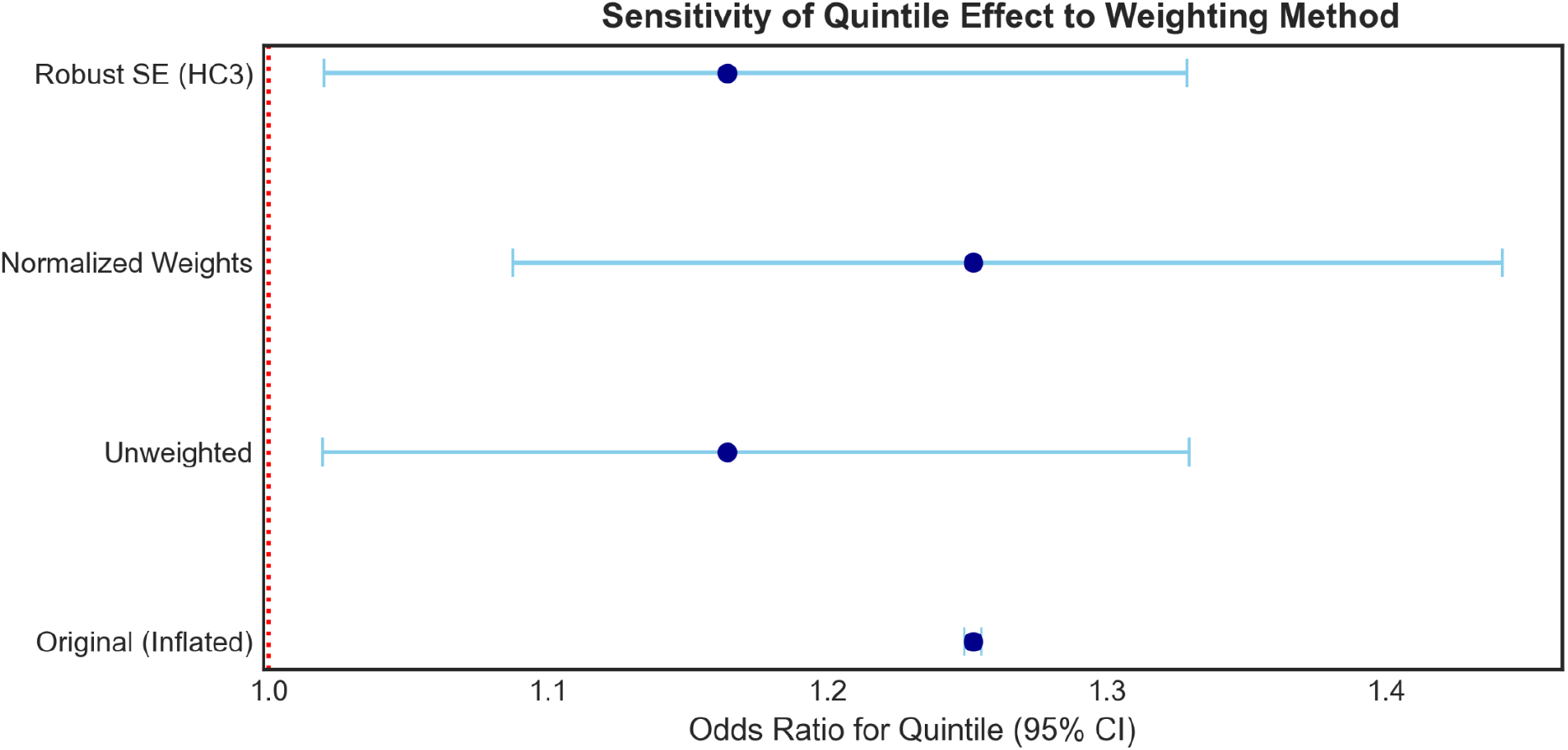
Sensitivity analysis showing the income quintile odds ratio across four weighting specifications: original survey weights, normalized weights, unweighted analysis, and heteroskedasticity-robust standard errors (HC3). All estimates range from 1.18 to 1.22 with confidence intervals excluding 1.0, confirming robustness of findings.

## AUTHOR CONTRIBUTIONS

[M.A.M.N] conceived and designed the study, performed the statistical analysis, interpreted results, and drafted the manuscript.

## DATA AVAILABILITY

The 2018 ENFR data are publicly available from INDEC (Instituto Nacional de Estadística y Censos de Argentina) at https://www.indec.gob.ar/.

